# Mendelian randomisation analysis using GWAS and eQTL data to investigate the relationship between chronotype and neuropsychiatric disorders and their molecular basis

**DOI:** 10.1101/2023.04.19.23288809

**Authors:** Shane Crinion, Cathy A. Wyse, Gary Donohoe, Lorna M. Lopez, Derek W. Morris

## Abstract

A wide range of comorbidities have been observed with neuropsychiatric disorders, of which sleep disturbances are one of the most common. Chronotype, a self-reported measurement of an individual’s preference for earlier or later sleep timing, is a proxy sleep measure that has been linked to neuropsychiatric disorders (NPDs). By investigating how chronotype influences risk for neuropsychiatric disorders and, vice versa, how risk for neuropsychiatric disorders influences chronotype, we may identify modifiable risk factors for each phenotype. By investigating the specific genetic mechanisms that are common to the risk of evening chronotype and the risk of NPDs, we may gain further understanding of the relationship and causal direction in these phenotypes. Here we use Mendelian randomisation (MR), a method used to explore causal effects, to 1) study the causal relationships between neuropsychiatric disorders and chronotype and 2) characterise the genetic components of these phenotypes. Firstly, we investigated if a causal role exists between six neuropsychiatric disorders and chronotype using the largest genome-wide association studies (GWAS) available. Secondly, we integrated data from expression quantitative trait loci (eQTLs) to investigate the role of gene expression alterations on these phenotypes. We also used colocalization to validate that the same variant is causal for gene expression and each outcome. We identified that the evening chronotype is causal for increased risk of schizophrenia and autism spectrum disorder and, in the opposite direction, that insomnia and schizophrenia are causal for a tendency towards evening chronotype. We identified twelve eQTLs where gene expression changes in brain or blood were causal for one of the tested phenotypes (bipolar disorder, chronotype and schizophrenia). These findings provide important evidence for the complex, bidirectional relationship that exists between these sleep and neuropsychiatric disorders, and use gene expression data to identify causal roles for genes at associated loci.

**Author Summary:** Sleep disturbances are commonly observed features of neuropsychiatric disorders. Chronotype, a behavioural manifestation of an individual’s preference for early or late sleep timing i.e. morning chronotype means a preference for earlier sleep and wake times, has been used as a behavioural marker of underlying circadian function. Here, we used data from the largest genetic studies available to test the causal relationship between chronotype and risk for neuropsychiatric disorders. We found that individuals with the evening chronotype have greater risk for schizophrenia or autism spectrum disorder. In the other direction, we found insomnia or schizophrenia diagnosis is causal for a tendency towards evening chronotype. We searched for DNA variants that influence chronotype or risk for neuropsychiatric disorders through alterations of gene expression in blood and brain tissues. We found twelve DNA variants with a significant effect on chronotype or risk of either bipolar disorder or schizophrenia. These results demonstrate that sleep and neuropsychiatric disorders have a complex bidirectional relationship and that the causal role of some genes is due to variants that alter gene expression.

## Introduction

Neuropsychiatric disorders include the adult-onset disorders of bipolar disorder (BD), major depressive disorder (MDD) and schizophrenia (SZ), and the child-onset conditions of attention deficit hyperactivity disorder (ADHD) and autism spectrum disorder (ASD). These disorders are multifactorial, caused by polygenic inheritance of many genetic variants in combination with various environmental factors (1). Symptoms of NPD are varied and include changes in behaviour, brain function and cognition, that can be episodic or continuous and range from mild to severe. Diagnosis of NPD is based on clinical evaluation of highly variable changes in behaviour and the heterogeneous nature of these disorders along with the significant overlap between diagnoses makes the identification of risk factors and molecular mechanisms difficult.

Sleep-related disorders are one of the most common comorbidities with neuropsychiatric disorders and typically individuals with a neuropsychiatric diagnosis experience some degree of difficulty sleeping (2). Insomnia is classed as a neuropsychiatric disorder and a sleep disorder and is one of the most common comorbidities of each neuropsychiatric disorder listed above (3). Evidence from genetic studies support an association between chronotype and changes in risk for neuropsychiatric disorders (4). Chronotype is a behavioural manifestation of an individual’s underlying circadian rhythm e.g. having the morning chronotype means having a preference to wake up and go to sleep earlier in the 24 hour day while having the evening chronotype means to prefer to wake up and go to sleep later.

It is often time-consuming, expensive or unethical to conduct randomised controlled trials (RCT) that could estimate the causal role of risk factors for neuropsychiatric disorders. An RCT cannot be used to investigate the causal relationship between sleep disorders and NPD, as patients could not be randomly allocated to sleep disruption groups. In such cases, Mendelian randomisation (MR) is a powerful complementary approach that can be applied to estimate the causal effect of an exposure on an outcome using instrumental variants (IVs) (5). For example, genome-wide significant (GWS) SNPs for chronotype can be used as IVs to estimate the causal effect of chronotype (exposure) on risk for a neuropsychiatric disorder (outcome). MR can be applied bidirectionally, i.e., using GWS SNPs for neuropsychiatric disorders to estimate the causal effect of neuropsychiatric disorders on chronotype.

To investigate the complex relationship between sleep and risk for neuropsychiatric disorders, we apply MR here to investigate the role of the modifiable risk factor chronotype in risk for neuropsychiatric disorders and predict the contribution of the genetic component to lifetime risk for both phenotypes on each other. MR studies have identified a causal relationship between four neuropsychiatric disorders (ADHD, BD, MDD and SZ) and sleep-related phenotypes (3, 6, 7, 8) and, in the opposite direction, a causal relationship between five sleep-related phenotypes (daytime napping, insomnia, morning person, long sleep duration and sleep duration) and changes in risk for neuropsychiatric disorders (3, 4, 6, 7). Our study here builds on previous work by using the largest GWAS available to date to explore causal relationships between chronotype and neuropsychiatric disorders. Our study also implements the new MR Pleiotropy RESidual Sum and Outlier (MR-PRESSO) method, which uses simulation to detect and remove horizontal pleiotropy (9). This may identify new causal associations or provide additional evidence for the chronotype-neuropsychiatric disorder relationship.

MR analysis can be extended to study the biological mechanisms that are individually involved in chronotype and risk for neuropsychiatric disorders. In this approach, expression quantitative trait loci (eQTLs) are used as IVs to estimate the causal effect of gene expression (exposure) on genetically-based outcomes, e.g., eQTLs are used to estimate the causative effect of changes in gene expression (exposure) on risk for a neuropsychiatric disorder or chronotype (outcome).

MR and colocalization are used together as an approach to provide evidence for the association between a genetic variant and disease risk (10). Previously this method has been implemented to link changes in gene expression to psychiatric and neurodevelopmental disorders and identify potential drug targets (11).

Overall, this study had two major objectives. The first was to investigate the causal relationship between chronotype and neuropsychiatric disorders using MR with GWAS summary statistics. To do this we used GWAS-based IVs associated with chronotype to investigate its effect on risk of neuropsychiatric disorders (ADHD, ASD, BD, MDD, Insomnia and SZ) and we used GWAS-based IVs associated with neuropsychiatric disorders to investigate their effect on chronotype. The second objective was to investigate the causal relationship between genetic variants associated with changes in gene expression and chronotype and/or neuropsychiatric disorders using MR with eQTL and GWAS summary statistics. The eQTL data employed was from PsychENCODE (http://resource.psychencode.org/) and GTEx (https://gtexportal.org/home/) for a range of brain tissues (hippocampus, hypothalamus and prefrontal cortex) plus whole blood.

## Materials & Methods

These methods follow the guidelines outlined in Strengthening the Reporting of Observational Studies in Epidemiology using Mendelian Randomization (STROBE)-MR which was developed to assist researchers to report MR studies in a clear and transparent way (12). STROBE-MR provides guidelines to improve the quality and assessment of MR studies. The checklist includes 20 items, of which the first three items have been addressed in the Title and Introduction of this manuscript (1: indicating MR as the study design, 2: explaining the scientific background and rationale of the study and justifying use of MR, and 3: stating specific objectives). The remaining items are addressed in the Materials and Methods (items 4-9), Results (items 10-13) and Discussion (items 14-20). The complete checklist and the relevant text from the manuscript for each point is outlined in Supplementary Table 1 (for first objective; bi-directional MR study of chronotype and neuropsychiatric disorders) and Supplementary Table 2 (for second objective; MR study of changes in gene expression on chronotype and risk of neuropsychiatric disorders). The tables highlight the implementation of the STROBE-MR guidelines throughout the manuscript.

### Objective 1: Bi-directional two-sample MR analysis of chronotype and neuropsychiatric disorders

To investigate causal relationships between neuropsychiatric disorders and chronotype, we performed a bi-directional two-sample MR analysis using genetic variants for each trait from GWAS data. We obtained summary statistics from the Psychiatric Genomics Consortium (PGC) for six neuropsychiatric disorders (ADHD, ASD, BD, MDD, Insomnia and SZ) (3, 7, 13, 14, 15, 16) and from the UK Biobank for chronotype (4). Chronotype was selected due to it being the sleep-related phenotype that has been subjected to the largest GWAS in terms of sample size and statistical power to detect associations of small effect. A description of the GWAS data used for neuropsychiatric disorders and chronotype is available in Supplementary Table 3.

To maximise power, we used the largest GWAS available for each phenotype. For each neuropsychiatric disorder, the individuals were classified as cases and controls based on the reported diagnoses in each GWAS. To prevent bias due to variation in the underlying sample population, all individuals included in these GWAS were of European ancestry. For SZ, the PGC had performed a multi-ancestry integration analysis (80% European + 20% East Asian; 14) so we obtained summary statistics for an entirely European ancestry sample. To avoid sample overlap, GWAS summary statistics that excluded UK Biobank samples were used, when available. Data for 403,195 participants was available from the UK Biobank for the morning chronotype. The measurement of chronotype refers to self-reported “morning person” as identified through the UK Biobank (n=449,734). A case for the morning person chronotype is a participant who, for the question “*Do you consider yourself to be:*”, answered “*Definitely a morning person*” or “*More a morning than an evening person*” whereas a control is a participant who answered with “*Definitely an ‘evening’ person*” and “*More an ‘evening’ than a ‘morning’ person*” in the UK Biobank data (4). All included studies reported a SNP as GWS if it reached the significance threshold (p < 5×10^-8^) and each study performed LD clumping on results to identify independent GWS loci (*r^2^* < 0.1). The range of clumping distance between SNPs in each of the selected GWAS ranged from 250 −3,000 kb (Supplementary Table 3).

During MR analysis we used genetic variants as a proxy for a trait and therefore it is important to ensure that the selected IVs are high quality and can accurately predict causality. There are three core IV assumptions that generally must hold in order for the MR study to be valid: 1) IVs are robustly associated with the risk factor; the relevance assumption. 2) IVs are not associated with any confounders; the independence assumption. 3) IVs have no association with the outcome that is not mediated through the risk factor; the exclusion restriction assumption (17). We address the relevance assumption by only using genetic instruments that were identified as GWS loci for each phenotype, providing evidence of a strong association with the phenotype.

We address the independence and exclusion restriction assumptions by using several MR methods to estimate causal effect and correct for bias due to horizontal pleiotropy. Inverse-variance weighted (IVW) meta-analysis was selected as the primary analysis for causal estimate due to its ability to provide maximum power possible. This method combines effect estimates for all genetic instruments to provide the greatest valid causal estimates, under the assumption of balanced pleiotropy (17). IVW MR was implemented using the mr_ivw function from the TwoSampleMR R package on the default settings (18). By default, the IVW random-effects model is performed, allowing overdispersion in the weighted linear regression (the residual standard error is not fixed to be one, but is not allowed to take values below one). We also selected that a minimum of four IVs were required to ensure a robust causal estimate was being produced in the MR model. We utilised the MR-PRESSO test to identify and remove outliers via a simulation approach that uses observed and expected distributions of tested variants to detect horizontal pleiotropy (9).

We additionally used the MR-PRESSO global test, which detects bias among all IVs by comparing the observed distance of all variants to the regression line, to detect horizontal pleiotropy and the MR-PRESSO distortion test to test for significant distortion before correction (9). These functions were implemented using the mr_presso function from the R package MRPRESSO. We also implemented a Bonferroni significance threshold to correct for multiple-testing (i.e., P < 0.05/12 tests for bidirectional analysis of 6 phenotypes).

Causal estimates were also made using MR-Egger regression (19), which corrects for average pleiotropic effects, and median-based estimate (20), which assumes >50% of instruments are valid. The effect estimates from these methods were compared to results from IVW. The MR-Egger intercept test was also used to estimate bias caused by horizontal pleiotropy (19). Heterogeneity was also measured using Cochran’s Q statistic (21). The additional MR methods were also implemented using the TwoSampleMR R package on the default settings. A number of plots were also generated to evaluate the degree of causal effects and identify bias. Forest plots were generated to visualise and compare causal odds ratio estimates and confidence intervals for each exposure and outcome pair identified. Scatter plots were also generated which plots the SNP-outcome association against the SNP-exposure association to visualise the causal estimate obtained from the MR analysis (22).

We extended the analysis to examine multiple exposures concurrently to further evaluate the degree of bias that may be introduced in causal estimates due to genetically similar phenotypes. We performed a multi-trait MR, which is implemented to correct when IVs for exposure 1 (E_1_) also have a significant effect on additional exposures (E_2_ → E_n_). The IVs for E_1_ are selected and the effect estimates are extracted for these IVs from E_n_ and integrated into the MR model. Due to prior knowledge of the significant genetic similarities between the psychiatric disorders (e.g. BD, MDD and SZ) (23), we employed multi-trait MR to investigate the effect of bias due to genetic similarities on each psychiatric disorder. We assessed the role of psychiatric disorders (SZ, BD and MDD) on chronotype by performing three multi-trait MR analyses, to assess each psychiatric disorder assigned as E_1_ and extract the corresponding IVs to test the effect on chronotype, i.e., MR analysis to investigate the effect of schizophrenia (E_1_=SZ, E_2_=BD and E_3_=MDD) on chronotype. We did not perform multi-trait MR for ADHD and ASD due to the low number of instrumental variants available for analyses. This was also implemented using the MR-PRESSO R package (9). We also utilised results from genetic correlation studies, generated using GWASatlas, to assess the direction of effect and to compare to results from MR (24).

We performed the MR analysis using the R version 4.0.2 (The R Foundation of Statistical Computing). The software used to perform this MR analysis was the R packages TwoSampleMR (https://github.com/MRCIEU/TwoSampleMR) and MR-PRESSO (https://github.com/rondolab/MR-PRESSO) and all plots were generated using the R packages ggplot2 (https://ggplot2.tidyverse.org/) and TwoSampleMR.

### Objective 2: MR analysis to assess the role of SNPs associated with changes in gene expression as risk factors for neuropsychiatric disorders and chronotype

This analysis uses MR to identify gene expression alterations that contribute to chronotype and/or neuropsychiatric disorders. This analysis was unidirectional and investigated the effect of gene expression (exposure) on neuropsychiatric disorders and chronotype (outcome) by integrating eQTL data with GWAS data within a MR framework to investigate the genetic basis of both neuropsychiatric disorders and chronotype, including overlap between them. Each phenotype was investigated separately with the aim of contributing to current knowledge on the genetic basis of each phenotype, validating previous findings from GWAS and other genomic analysis methods, and/or finding previously unidentified variants or genes that link neuropsychiatric disorders and chronotype.

To strengthen the evidence of a causal relationship, we followed up the MR analysis by performing colocalization on the top findings. MR and colocalization can be used together to validate that the variant in a genomic region has been identified as causal for both the exposure and the outcome through MR. In this analysis, MR assessed the association of an exposure (eQTLs) with an outcome (neuropsychiatric disorders and/or chronotype). Evidence of an association indicated that the eQTLs were causal for gene expression alterations that contribute to the outcome phenotype. Colocalization was then performed where strong evidence for an association between an exposure and outcome is found through MR and assessed whether each is caused by the same or distinct variants (10). This analysis was motivated by the approach taken by Baird et al. (2021) to perform two-sample MR using eQTLs as instruments to predict effects on neuro-phenotypes (11)

We used eQTL summary statistics from blood and the brain. We selected brain regions based on their relevance in the context of neuropsychiatric disorders and sleep phenotypes such as a overlapping relevance to both phenotypes (Supplementary Table 4). Prefrontal cortex data from PsychENCODE data was included as the largest dataset of brain eQTLs available (n=1387), while GTEx whole blood was included as the largest dataset of eQTLs from GTEx (n=670) and as a proxy for brain tissue. The specific brain regions that were selected were based on their relevance to neuropsychiatric disorders, circadian rhythm or both and these were cortex (n=205), hippocampus (n=165) and hypothalamus (n=170).

For each of the selected tissues, we obtained gene expression data from cis-eQTL summary statistics provided by the Genotype-Tissue Expression (GTEx) project version 8 (https://gtexportal.org/) (25) and the PsychENCODE consortium (https://psychencode.synapse.org/) (26). The number of samples per tissue is outlined in **Supplementary Table 4** and ranged from 165 samples (GTEx, hippocampus) to 1,387 samples (PsychENCODE, prefrontal cortex). Information on the measurement, QC and selection of genetic variants is available in the respective studies.

Study participants were primarily of European ancestry. eQTL summary statistics from PsychENCODE were generated using data from samples of European ancestry. eQTL summary statistics from GTEx consisted of 84.6% white, 12.9% African American and 1.3% Asian participants.

Similar to the first analysis, we used the beta coefficient to report the strength of the effect of the exposure (gene expression from eQTL summary statistics) on each of the seven outcomes (six neuropsychiatric disorders and chronotype) individually. The IVs for gene expression were selected by using a statistical significance threshold of p < 5 x 10^-8^ and LD clumping was performed by the clump_data function from the TwoSampleMR R package on the default settings, to obtain a set of independent eQTL instruments. SNPs were also filtered to include only those available in the 1000 Genomes EUR population with r^2^ > 0.001 and all SNPs within clumping distance of 10,000kb window of the top hit were removed. This resulted in a total of 4,374 unique SNPs and 6,315 genes to study using MR analysis (Supplementary Table 4). As in our first analysis, we implemented the MR analyses using the TwoSampleMR R package, using the mr_wald_ratio function.

The causal estimate was obtained using the single instrument Wald ratio, that divides the gene-outcome association by the gene-exposure association (27). Similarly to the analysis performed using the SMR package, developed by <u>Zhu et al.</u> (2016), we apply MR to assess gene expression on complex traits and exploit the Wald ratio in conjunction with a colocalization method (28). Colocalization analysis was performed to support evidence from MR where a statistically significant causal effect was found between an eQTL and a neuropsychiatric disorder and/or chronotype. Colocalization analysis validates that the same eQTLs are causal for changes in gene expression and the neuropsychiatric disorder and/or chronotype. A non-zero result from colocalization analysis indicates that the same variants are causal in both GWASs and eQTL studies, due to either causality or pleiotropy. We performed colocalization using the coloc.abf function from the coloc R package (29). This function calculates posterior probabilities (PP) using Bayes factors to assess the association of a SNP with two traits and calculates a PP for four hypotheses, of which hypothesis 4 (PP4) > 0.7 is considered evidence that a common causal variant is found between the two traits. We also undertook to validate the IV assumptions by only using eQTL instruments found to have a strong effect on the exposure, based on the statistical significance threshold of P-value < 5×10^-8^.

We calculated a beta effect size, standard error and p-value from each Wald test to test the null hypothesis that gene expression alterations were not correlated with risk of neuropsychiatric disorders and/or chronotype. The test for causal effect from MR analysis showed a statistically significant association if the statistical result passed the Bonferroni correction threshold of p < 0.05/7 (correcting for testing of six neuropsychiatric disorders plus chronotype).

## Results

### Bi-directional two-sample MR analysis of chronotype and neuropsychiatric disorders

#### Causal effects of chronotype on neuropsychiatric disorders

Our two-sample MR analysis demonstrated that the morning chronotype is causal for lower risk of two neuropsychiatric disorders (**Figure 1**). This includes an association between morning chronotype and reduced risk for ASD (OR = 0.88, CI = 0.82-0.94, IVW P = 4.37×10^-4^) and reduced risk for schizophrenia (OR = 0.91, CI = 0.95-0.97, IVW P = 2.19×10^-3^). We also detected an association between morning chronotype and reduced risk for MDD (OR = 0.95, CI = 0.91-0.99, IVW P = 1.53×10^-2^), however the estimate was not statistically significant after correcting for multiple tests (Supplementary Table 5).

**Figure 1.**
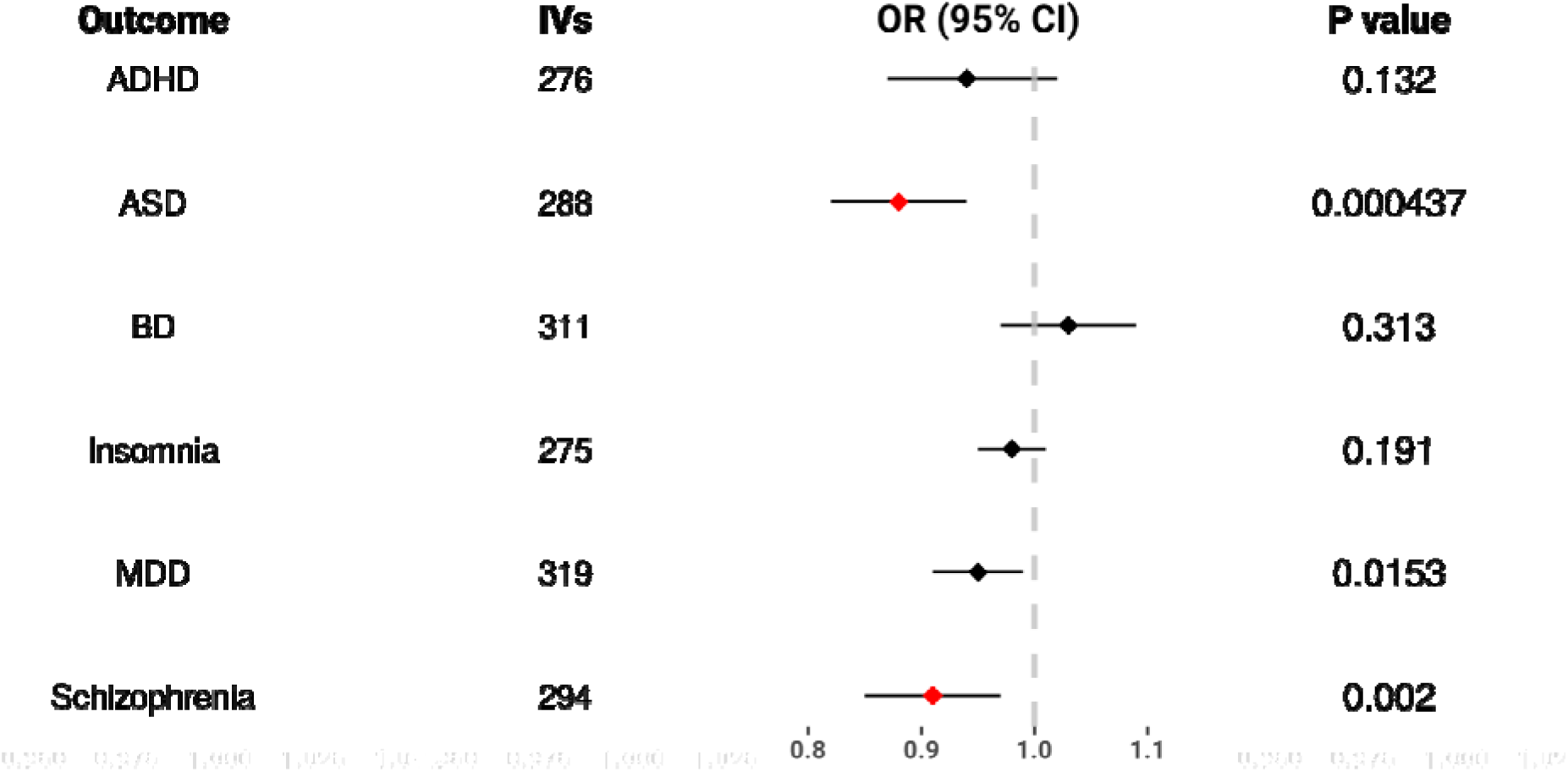
MR forest plot of chronotype (exposure) vs. neuropsychiatric disorder (outcomes). Plot showing MR IVW meta-analysis effect estimates (OR). OR above 1 indicate a tendency towards the morning chronotype and OR below OR indicate a tendency towards the evening chronotype. Each line represents CI and red dots identify associations that reach the Bonferroni-corrected significance threshold. ADHD, attention-deficit hyperactivity disorder; ASD, autism spectrum disorder; BD, bipolar disorder; CI, confidence intervals; IVs, instrumental variants (#GWS SNPs for the exposure (chronotype)); MDD, major depressive disorder.

To investigate the accuracy of the MR IVW method for causal effect estimates, a series of additional MR analyses were performed. We calculated causal effect estimates using MR-PRESSO, MR Egger and the median-based MR estimates and by comparing across methods, we found similar beta effect sizes albeit with wider confidence intervals (Supplementary Table 5). **Figure 2** shows that there is consistency across numerous MR methods in terms of effect size and direction for the two statistically significant results, ASD (**Figure 2a**) and SZ (**Figure 2b**). We also performed IV outlier removal by using the MR-PRESSO outlier test which determines which IVs may be confounding results by a simulation approach. The results indicated that 27 outliers for SZ and no outliers for ASD were observed and subsequently these were removed from the MR analysis, resulting in stronger associations identified between the significant exposure-outcome pairs.

**Figure 2.**
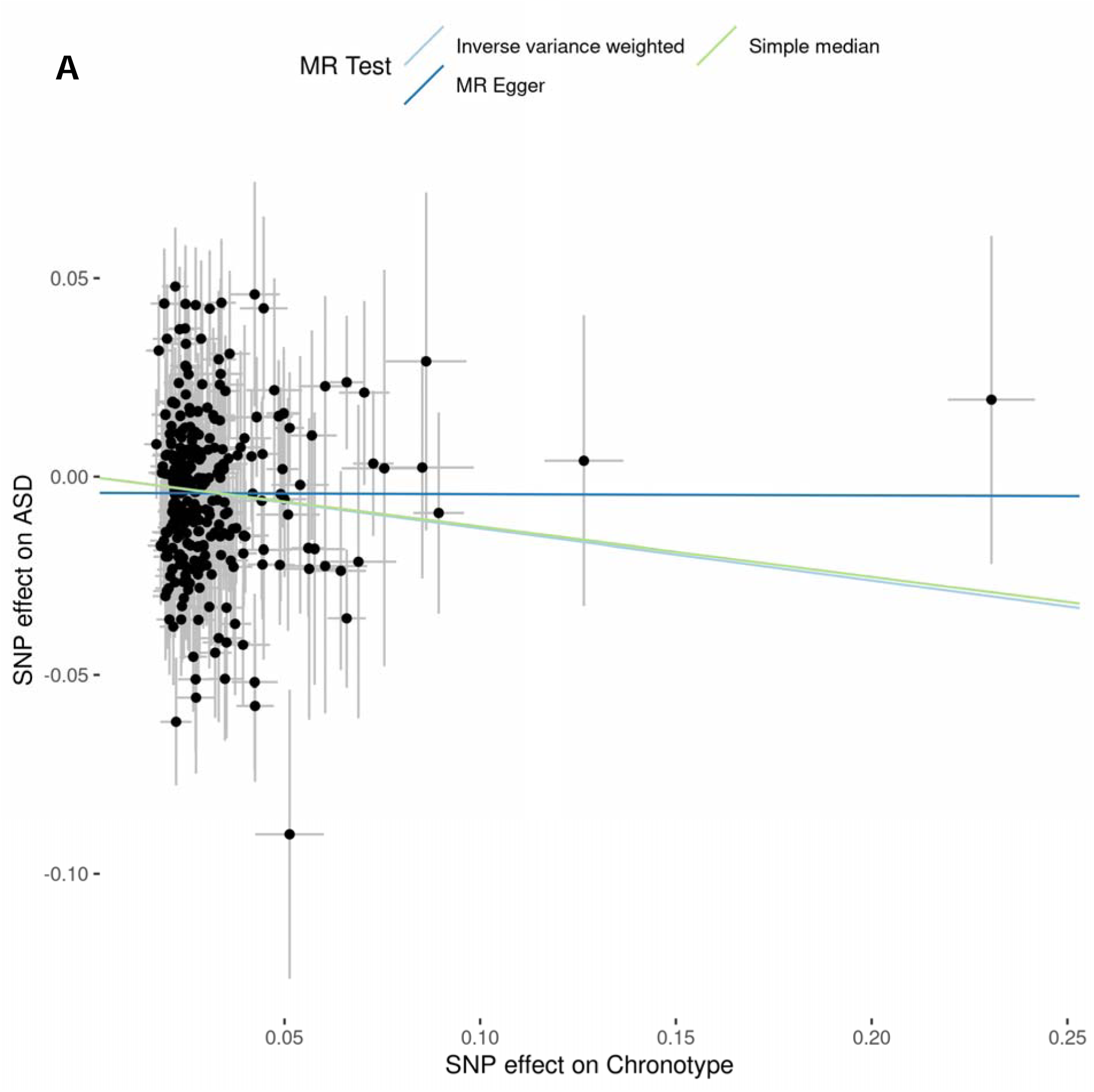
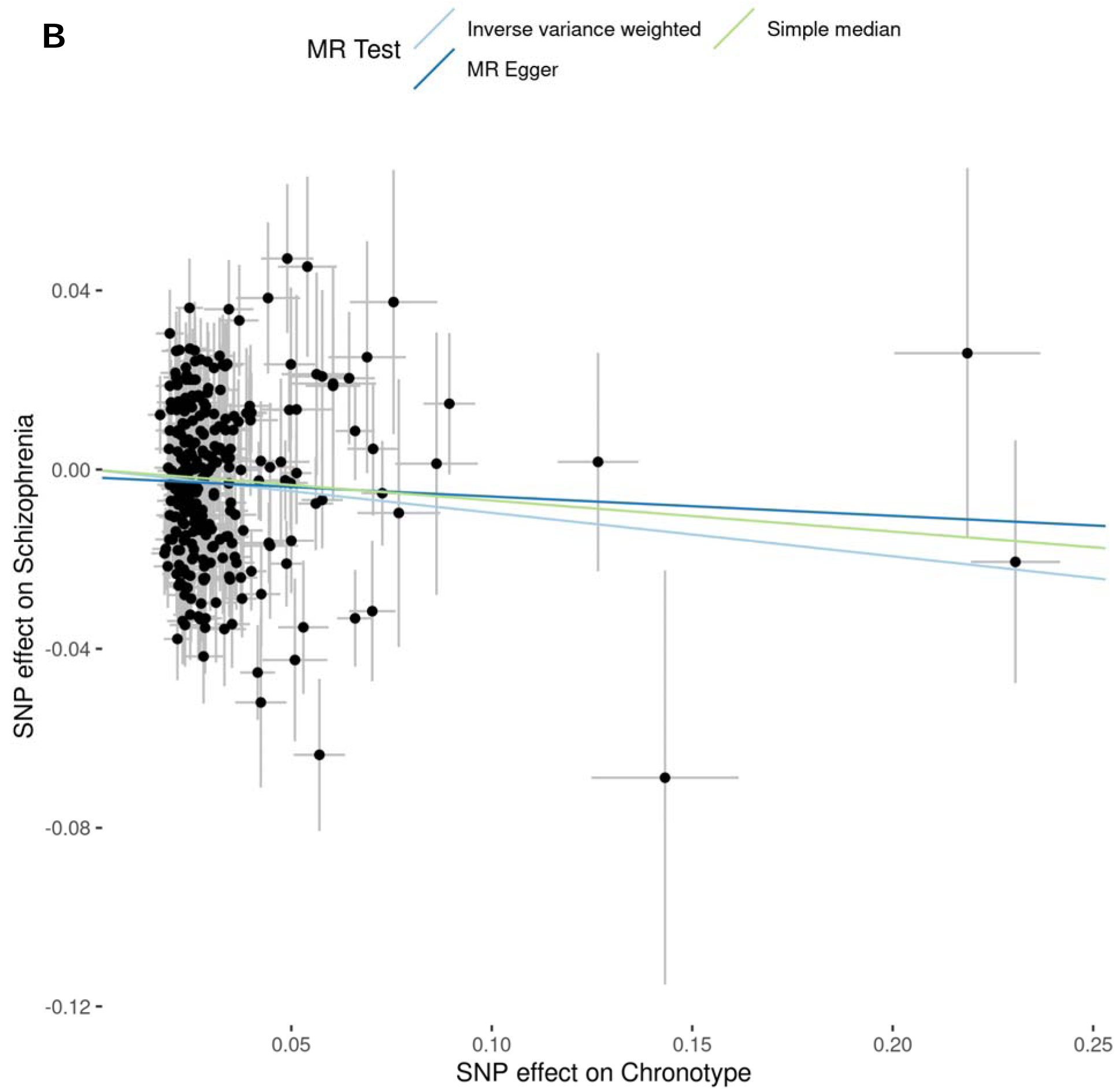
MR scatter plot of chronotype (exposure) vs neuropsychiatric disorders risk (outcome). This plot shows the effect size comparison (beta) for each instrumental variant used in the MR analysis. The x-axis indicates effect estimate for each SNP on morning chronotype while the y-axis indicates the effect estimate for the SNP on ASD (**Figure 2A**) and SZ (**Figure 2B**). Error bars at each point indicate the standard error for each IV in a single-variant MR analysis. The slopes from each coloured line represents the results from meta-analysis across multiple MR methods performed and the slope represents the degree of association.

#### Causal effects of neuropsychiatric disorders on chronotype

Our results of two-sample MR analysis demonstrated a greater genetic risk for insomnia (OR = 0.95, CI = 0.92-0.95 and IVW P = 8.92×10^-6^) and SZ (OR = 0.88, CI = 0.82-0.94, IVW P = 5×10^-4^) is causal for tendency towards the evening chronotype (**Figure 3**). We calculated causal effect estimates using MR-PRESSO, MR Egger and the median-based estimate and by comparing across methods. We found similar beta effect sizes albeit with wider confidence intervals (Supplementary Table 5). **Figure 4** shows that there is consistency across the various MR methods performed for effect size and direction for the two statistically significant results, insomnia (**Figure 4a**) and SZ (**Figure 4b**). We also performed IV outlier removal by using the MR-PRESSO outlier test. The results indicated that 24 outliers for insomnia and 14 outliers for SZ were observed and subsequently these were removed from the MR analysis (Supplementary Table 5), resulting in stronger associations identified between the significant exposure-outcome pairs.

**Figure 3.**
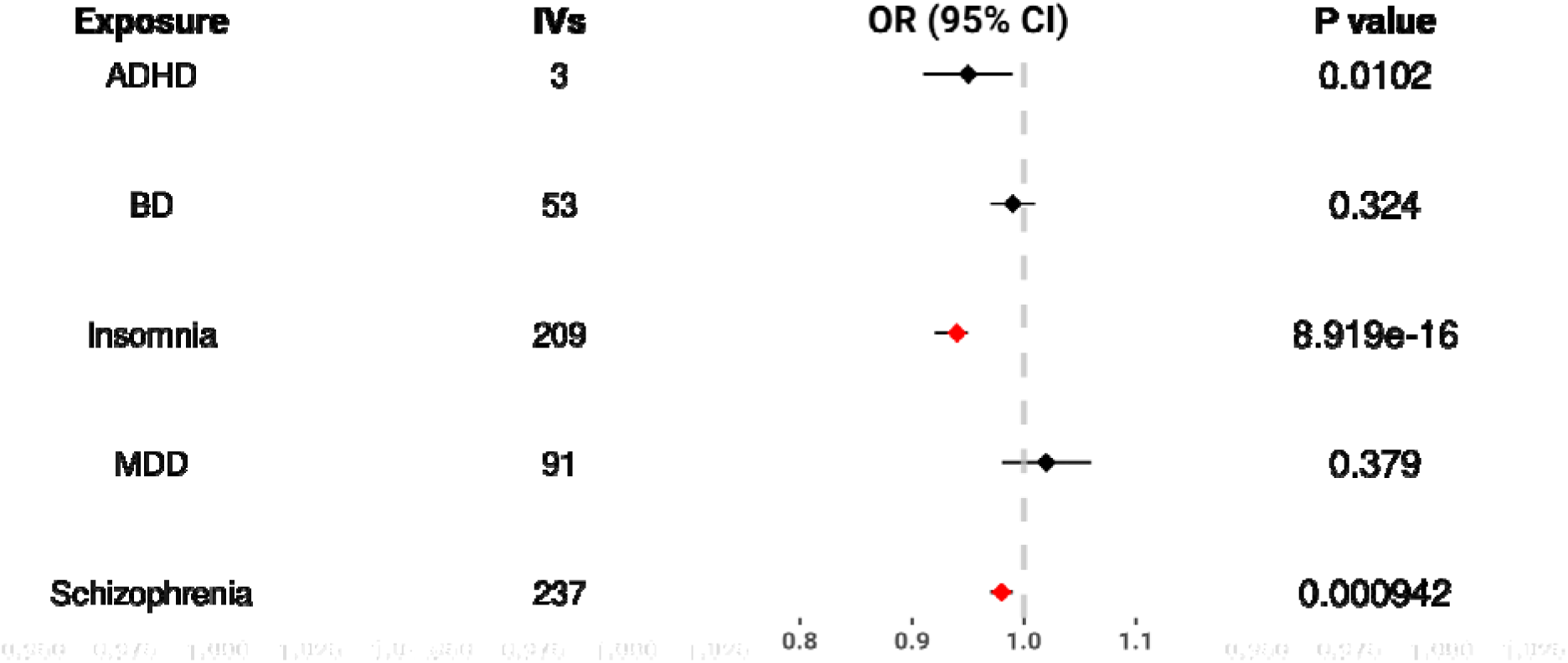
MR forest plot of neuropsychiatric disorder (exposure) vs. chronotype (outcome). Plot showing MR IVW meta-analysis effect estimates (OR). OR above 1 indicate a tendency towards the morning chronotype and OR below OR indicate a tendency towards the evening chronotype. Each line represents CI and red point colours identify associations that reach the Bonferroni-corrected significance threshold. ADHD, attention-deficit hyperactivity disorder; BD, bipolar disorder; CI, confidence intervals; IVs, instrumental variants (#GWS SNPs for the exposures (fiv neuropsychiatric disorders)); MDD, major depressive disorder.

**Figure 4.**
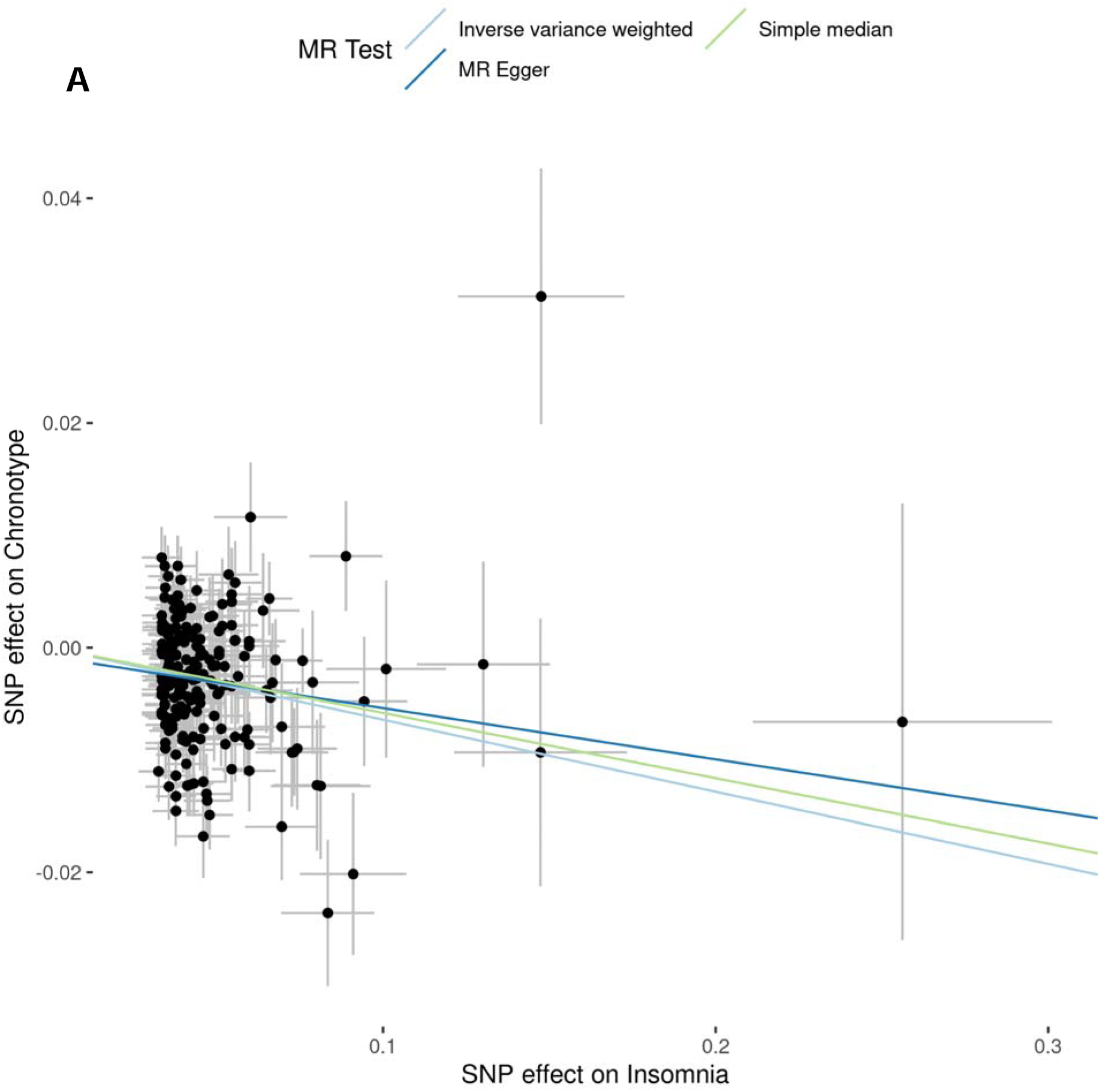
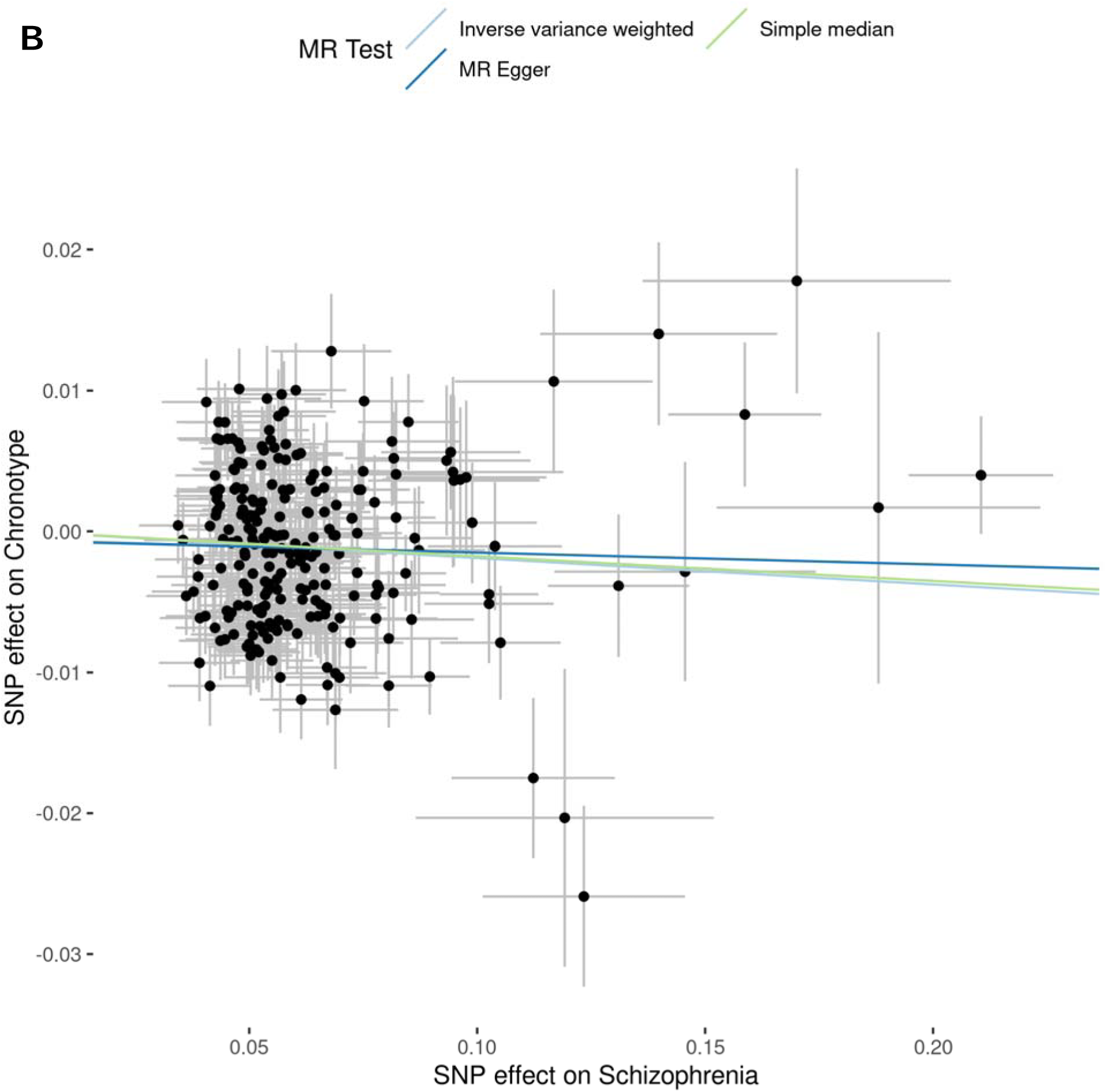
MR scatter plot of neuropsychiatric disorders risk (exposure) vs chronotype (outcome). This plot shows the effect size comparison (beta) for each instrumental variant used in the MR analysis. The x-axis indicates the effect estimate for each SNP on insomnia (**Figure 4a**) and SZ (**Figure 4b**) while the y-axis indicates the effect estimate for the SNP on the morning chronotype. Error bars at each point indicate the standard error for each IV in a single-variant MR analysis. The slopes from each coloured line represents the results from meta-analysis across multiple MR methods performed and the slope represents the degree of association.

A similar pattern of results was observed for a number of sensitivity tests in either direction of MR analysis. Cochran’s Q statistic found evidence for SNP effect sizes heterogeneity however we showed through consistency in the causal effect and direction, as predicted in sensitivity tests, it is unlikely to introduce bias from horizontal pleiotropy. As we are using MR for complex traits, which consist of many SNPs with very small effect sizes across numerous genes and biological functions, we expect that many of the SNPs used as IVs will have wide variability in their estimated effect size. We also found no significant results from the MR Egger intercept test (p > 0.05), suggesting that no bias due to horizontal pleiotropy was observed. This is also supported by the individual SNP tests (leave-one-out analyses), where no individual SNP-exposure association was found to dominate the estimate of the causal effect (30). Full details on the sensitivity tests of the bidirectional MR analyses are available in Supplementary Table 5.

The results from multi-trait MR also show good support for these findings. We assessed the causal effect of psychiatric disorders (BD, MDD and SZ) on chronotype using three MR analysis designs and found that the predicted effect sizes and standard errors correlated with those generated in the uni-trait MR analyses. Statistical significance also remained for the effect of SZ on chronotype and a number of outlier IVs were no longer removed, due to accounting for the bias introduced from BD and MDD (Supplementary Table 6).

We also compared the results of MR analysis with results from genetic correlation, generated using GWASatlas. We verified that similar results were obtained using genetic correlations and showed that the direction of effect is in line with our results, indicating that chronotype is negatively correlated with risk of all the neuropsychiatric disorders that we investigated using MR. A similar pattern was found where a significant genetic correlation was found between chronotype and ASD, MDD and SZ (Supplementary Figure 1).

Finally, in order to assess the effect of heterogeneity due to sample population variation, we calculated causal effects between chronotype and a mixed ancestry SZ sample (20% East Asian + 80% European) and demonstrated that similar effect estimates were obtained by comparing the results from the European ancestry study (Supplementary Table 4).

#### MR analysis to assess the role of SNPs associated with changes in gene expression as risk factors for neuropsychiatric disorders and chronotype

We analysed the effect of changes in gene expression on outcomes by performing two-sample MR using beta effect estimates for IVs for gene expression from eQTL summary statistics and extracting beta effect sizes for neuropsychiatric disorders and chronotype from GWAS data (Supplementary Table 7). We used a Wald ratio to estimate causal effects of 36,530 eQTL-gene pairs, based on the number of unique eQTL-gene pairs passing LD clumping that had a statistically significant association in blood or brain tissue. Bonferroni correction was performed by tissue specific thresholds (outlined in Supplementary Table 8) that were used to determine the significance of the causal effects found through MR and revealed 175 significant causal effects where an outcome was mediated through gene expression alterations. These results were from 66 unique eQTLs across 161 unique genes (Supplementary Table 7).

We found a significant impact of gene expression alterations, mediated through eQTLs, in numerous genes for all of the outcomes that we analysed (ASD: 10 genes, ADHD: 2, BD: 34, Insomnia: 2, MDD: 2, SZ: 105 and chronotype: 20, Supplementary Table 8). We then used colocalization, an established method used to provide complementary evidence to predict causal relationships between gene expression and complex traits, and found that strong evidence for a causal effect remained for twelve of these associations, with PP4 > 0.7 (Supplementary Table 7). **Figure 5** shows these twelve eQTLs with a significant causal role in neuropsychiatric disorders or chronotype. Supplementary Table 7 includes the effect estimates (β) and statistical significance (P) measured for each instrumental variant in the original eQTL (β_exp_ and P_exp_) and GWAS (β_out_ and P_out_).

**Figure 5.**
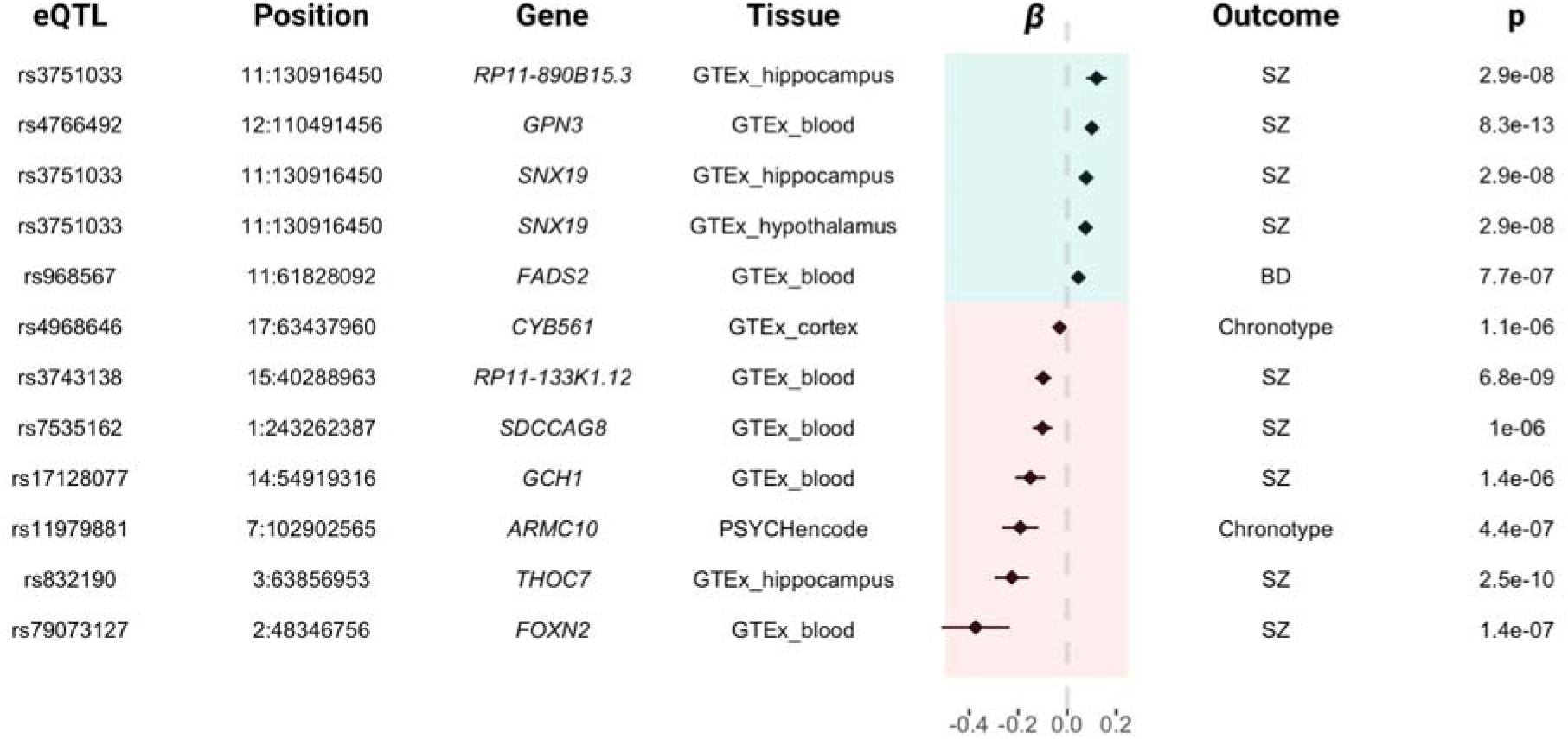
Forest plot for significant associations found from MR analysis and colocalization of the causal effect of gene expression on chronotype or neuropsychiatric disorders. For each eQTL SNP, the beta (β) represents the effect size of gene expression alterations in the respective gene for the corresponding outcome and tissue. The point an lines at each row represent the strength of effect and confidence intervals for each comparison. The vertical dashed lin represents beta = 0. Results with beta shaded in blue indicate that increased gene expression is associated with the increased risk for neuropsychiatric disorders or increased tendency for the morning chronotype. Results with beta shaded in re indicate that decreased gene expression is associated with increased risk for the phenotype. The p-value indicates the strength of the effect on the outcome and the tissue column indicates the source of the eQTL. Each result passed the MR test for causal effect estimate and colocalization for the same causal variant.

Six of twelve associations were blood-mediated and nine of twelve associations were for SZ. It is notable that for the rs3751033 eQTL, we demonstrated a pleiotropic effect on *RP11-890B15* in the hippocampus and on *SNX19* in the hippocampus and hypothalamus (**Figure 5**).

As we were interested in finding supporting evidence for the causal effects of the twelve eQTLs, we investigated if the associated eQTLs or genes have been previously linked to neuropsychiatric disorders and/or chronotype. *CYB561* was not previously implicated in chronotype. The result for rs4968646 and chronotype indicates that the causal effect at this locus can be attributed to altered expression of *CYB561* and not the *TANC2* gene, which has previously been reported to contain a SNP (rs17682747) that is in high LD with rs4968646 and associated with chronotype (31). MR analysis provided further evidence for the association of *RP111-890B15*, *GPN3* and *SNX19* with SZ, and *FADS2* with BD. For rs4968646, rs17128077 and rs79073127, no published evidence was found for the association of an eQTL/gene with the phenotype however results from PheWAS analysis, accessed through Open Targets Genetics, show evidence of an association between numerous eQTLs and phenotypes (Supplementary Table 9). Other eQTLs (rs3751033, rs4766492, rs832190, rs79073127) were previously reported in the literature and MR provides further validation of these associations here.

The tool LDlink proved useful for investigating the phenotypes that were linked to a SNP by proxy through a SNP in high LD. In most cases, the eQTL that was identified as causal through MR analysis, for a given phenotype, was in high LD with a SNP that was previously linked to that phenotype. For one eQTL (rs11979881 in *ARMC10* which was linked to chronotype through MR), neither the SNP or any SNP in high LD was previously linked to chronotype, indicating that we may have identified new evidence for the link between the eQTL rs11989881 and gene *ARMC10*. MR analysis also validates previously known links between sleep phenotypes and neuropsychiatric disorders (*FOXN2, GCH1, CYB561*). More details on each of the findings and a conclusion on the importance of each eQTL can be found in the Supplementary Table 9.

## Discussion

We identified a causal effect of chronotype on risk for SZ and ASD, indicating that an individual with the morning chronotype is at lower risk of developing either disorder. <u>Jones et al.</u> (2019) and Sun et al. (2019) did previously report a causal effect of chronotype on SZ but that was using a smaller sized SZ GWAS dataset (4, 32). Our finding of a causal effect of chronotype on ASD has not been previously reported through MR but the direction of effect is consistent with previous findings using measures of genetic correlation (14). In the opposite direction, we found that insomnia and SZ both have a causal effect on chronotype, i.e., a genetic predisposition to insomnia or SZ is causal for tendency towards the evening chronotype. The causal effect of insomnia on chronotype is consistent with observations in individuals with insomnia i.e. those with insomnia will typically wake and sleep later. These findings align widely with findings from other studies on the relationship between chronotype and neuropsychiatric disorders, whereby the evening chronotype is linked to increased risk and the morning chronotype is linked to lower risk for neuropsychiatric disorders (6, 7, 33). We built upon the analyses performed in previous studies by including the MR-PRESSO method, which allowed a new approach to identify and correct for outliers contributing to horizontal pleiotropy. In addition, we employed the MR-STROBE method to deliver a clear and transparent MR analysis.

The validity of these results was tested through a number of sensitivity tests, to assess that the fundamental assumptions of MR were met: 1) Relevance assumption: The instrumental variables were selected based on p-value, as reported in the respective GWAS; 2) Independence assumption and 3) Exclusion restriction: The causal effect estimates were also created using MR-Egger, weighted median and mode-based methods (19, 20). The effect estimates yielded were similar to those in the IVW method. The MR-Egger intercept was also used and found no evidence that results deviated from 0, indicating little evidence for directional genetic pleiotropy. Additionally, the multi-trait MR was used to account for confounding that may exist due to other existing neuropsychiatric disorders (BD and MDD), with overlapping features. These sensitivity tests gave sufficient evidence that MR analysis would produce reliable estimates for causal effects.

The bidirectional relationship between SZ and chronotype can be interpreted as the morning chronotype has a protective effect against SZ and, in the opposite direction, SZ can be a contributing factor in someone having the evening chronotype. This bidirectional causal relationship we identified between SZ and chronotype indicates that the evening chronotype can be both a cause and effect of the SZ. This suggests that there may be a type of positive feedback occurring whereby the disruption to sleep can initiate onset of SZ or perhaps make symptoms worse. An individual with SZ may also then experience further sleep disruptions which may lead to further disadvantageous symptoms of the disorder. The bidirectional relationship may be due to the heterogenous and polygenic nature of SZ and sleep-based phenotypes. Improved definition of sleep phenotypes and endophenotypes of SZ, and a better understanding of their genetic basis, are required to further assess the complex relationship between sleep and SZ.

Our second analysis was a unidirectional MR using eQTL and GWAS data, and was combined with colocalization to investigate the causal effects for alterations in gene expression on chronotype and six neuropsychiatric disorders. We found twelve instances where a unique eQTL-gene association was causal for either chronotype, BD or SZ. In some cases, these data support previously reported findings that have been identified through GWAS and PheWAS analyses such as the association of *SNX19* with SZ, which has been reported in multiple studies (Supplementary Table 9). In other cases, our data provides new evidence for the effect of SNP-mediated gene expression on risk of chronotype and neuropsychiatric disorders. The genes that have been identified may potentially inform on the biological mechanisms that could drive the genetic component of these phenotypes, e.g. intracellular vesicle trafficking is mediated by *SNX19* (34), and subsequently this process may be linked to SZ risk. Here we have used the MR framework to understand in part how gene expression alterations influence neuropsychiatric disorders and chronotype.

There are a number of strengths and weaknesses for each analysis. A major strength of the study is the application of the MR-STROBE method, which provided valuable guidance on the necessary information and additional analyses required to ensure robust analysis. For the first study, the framework was useful for planning, selecting the sensitivity tests and selecting figures and allowed us to perform analyses with reliable results through a proven method. We found that when applied to the second study the MR-STROBE method needed some modifications to be applicable in a multi-omic setting where additional methods such as colocalization are typically integrated into the analysis. Nevertheless, MR-STROBE remained useful for identifying the required study design components and methods to ensure that the second study was robust.

In our first analysis, we estimated causal effects using multiple sensitivity analyses (MR-PRESSO, MR Egger and median based estimates) to test for horizontal pleiotropy or bias due to potential confounders. We also made efforts to prevent bias due to sample heterogeneity by using only European samples and performing LD pruning to prevent correlation between SNPs used as genetic instruments. We utilised the MR-PRESSO outlier test to identify and remove pleiotropic outliers and ensure robustness of our study design. We also compared the results to those from studies of genetic correlation to validate that the directions of effect were consistent with our findings.

There are a number of limitations to the first analysis. While we have demonstrated significant evidence for a number of causal relationships, the biological basis of cause and effect remains to be elucidated. This relates to the difficulty in interpreting the results of MR studies when we use MR to study causal relationships of complex traits. When we perform MR using a single instrument with a clearly defined biological mechanism, we can design and guide treatments using this as a target. However due to the nature of complex traits, where we are dealing with many SNPs that have very low effect size, the biological impact of the individual SNPs is low and MR can only be used for epidemiological purposes.

Another limitation to this analysis is that we perform MR using only European ancestry samples. While two sample MR requires that the same underlying population is used, we believe that the development of multi-ancestry MR methods will be important to fully utilise newer GWAS that are beginning to include samples from numerous ancestry groups. Expansion of GWASs across diverse populations will lead to sample sizes that are large enough to detect SNPs that can be used as strong IVs in MR and identify new biological functions implicated in sleep and neuropsychiatric disorders. Finally, it is important to consider that chronotype is a self-reported measurement that may be biassed due to individual subjectivity and differences between individuals in terms of their stage of life and environment (35).

The second analysis had a number of strengths including integrating functional data from PsychENCODE and GTEx. The integration of eQTL data allows us to determine that the effect of a SNP on the phenotype is mediated through gene expression alterations. The method also indicates the degree of effect size, which can be valuable for prioritising genes. Another strength is that we have extended to integrating colocalization analysis, meaning that we find evidence to support the causal effect found through MR by coupling with evidence for the same causal variant implicated in the phenotype and gene expression. The results of colocalization demonstrated that, while a non-zero result can be obtained from MR, the result can lack evidence for a causal effect.

Further limitations of this analysis are that we used eQTLs detected in a relatively small number of brain samples and the decision to only include brain subregions that we deemed most relevant to our studied phenotypes. One other limitation found in this case is the lack of developmental stage-specific eQTLs, which may inform on varying effects of SNPs on gene expression levels during various stages of development. The effect sizes are also small and spread across numerous genes, meaning that feasibility for clinical application is limited. Finally, it is surprising that we did not achieve more significant findings from PsychENCODE, which was the largest source of eQTL data that we had available.

Overall, our MR analysis validated previous findings and provided new evidence for the biological aetiology of our neuropsychiatric disorders, chronotype and their complex relationship. We demonstrated that chronotype and neuropsychiatric disorders have a direct causal effect on each other, and that chronotype and risk for neuropsychiatric disorders (BD and SZ) are influenced by gene expression alterations in the blood and/or specific brain regions (hippocampus, cortex and hypothalamus). We identify a number of genes that are implicated in both chronotype and neuropsychiatric disorders that give important insight into the overlapping biology that is known to exist between neuropsychiatric disorders and sleep phenotypes. Future analyses may focus on diversifying the sources and types of GWAS and multi-omic data to integrate data on additional sleep phenotypes, regulatory function and brain regions that may mediate the links between neuropsychiatric disorders and chronotype. Integration of trans-eQTL data may provide further insights into the genetic basis of these phenotypes. Follow-up studies may also include a complete comparison to results from other methods that integrate expression data which include TWAS and S-PrediXcan. Interventions such as light therapy, which can manipulate an individual’s sleep timing (36), are available to influence chronotype. Given our study findings that evening chronotype is causally associated with risk of NPDs, these interventions may be useful to alter sleep timing towards the morning chronotype and reduce risk of NPDs.

## Bioinformatics Resources

The sources used in the literature search included GWAScatalog (37, https://www.ebi.ac.uk/gwas/home), Open Targets Genetics (38, https://genetics.opentargets.org), ClinVar (39, https://www.ncbi.nlm.nih.gov/clinvar), SNPedia (40, https://www.snpedia.com) and dbSNP (41, https://www.ncbi.nlm.nih.gov/projects/SNP).

## Data and data sharing

We deposited the full scripts for our MR analysis on GitHub (**Objective 1** available at shanecrinion/causal-inference and **Objective 2** available at shanecrinion/mendelian-randomisation-qtl).

## Data Availability

All data produced in the present work are contained in the manuscript

---

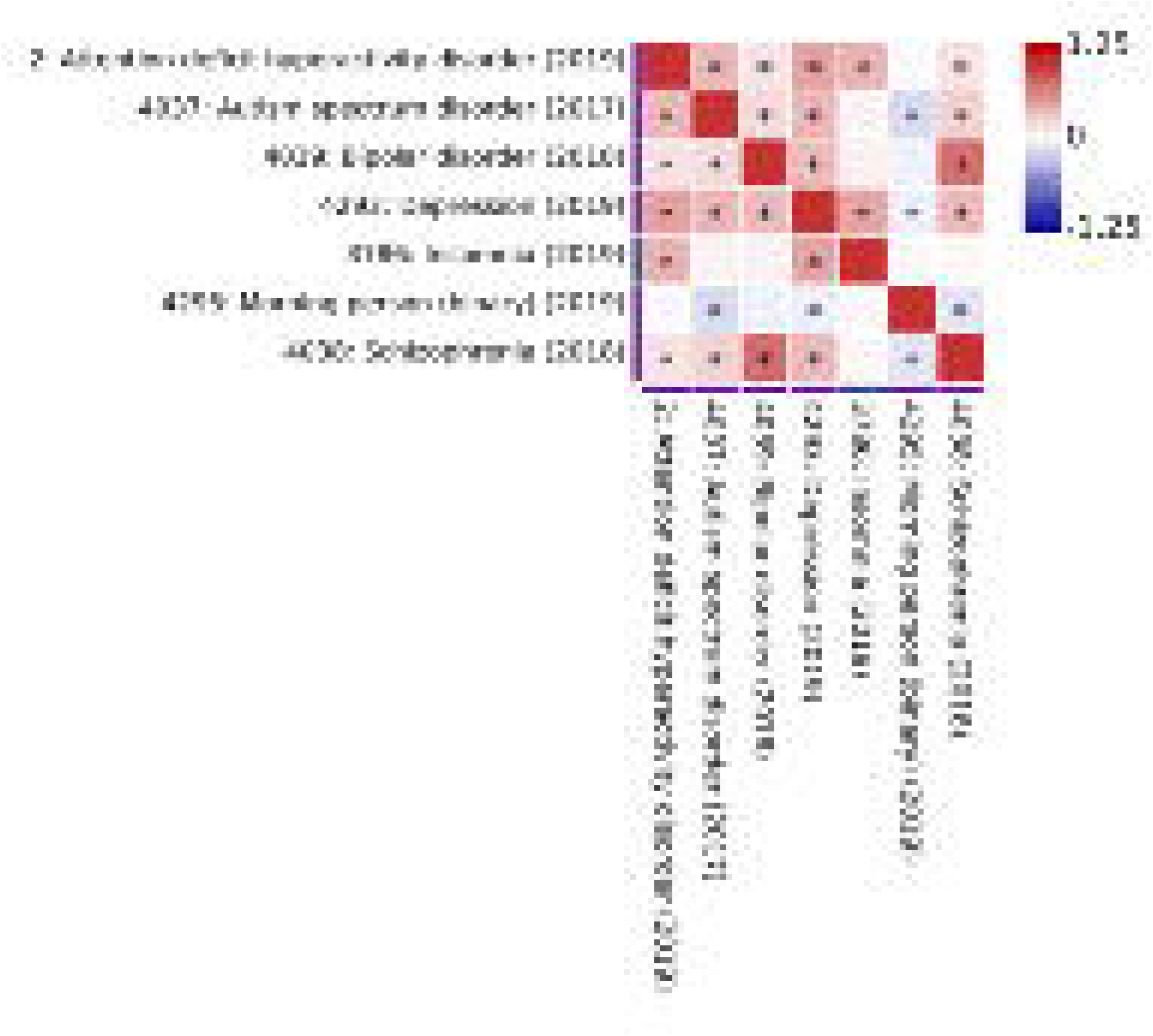

